# A Scoping Review of the Experience of Implementing Population Testing for SARS-CoV-2

**DOI:** 10.1101/2021.01.11.21249571

**Authors:** Clare R Foster, Fiona Campbell, Lindsay Blank, Anna Cantrell, Michelle Black, Andrew C K Lee

## Abstract

**Background:** The SARS-CoV-2 pandemic has led to the swift introduction of population testing programmes in many countries across the world, using testing modalities such as drive-through, walk-through, mobile and home visiting programmes. Here, we provide an overview of the literature describing the experience of implementing population testing for SARS-CoV-2.

**Methods:** We conducted a scoping review using Embase, Medline and the Cochrane Library in addition to a grey literature search. We identified indicators relevant to process, quality and resource outcomes related to each testing modality.

**Results:** 2,999 titles were identified from the academic literature and the grey literature search, of which 22 were relevant. Most studies were from the USA and the Republic of Korea. Drive-through testing centres were the most common testing modality evaluated and these provided a rapid method of testing whilst minimising resource use.

**Conclusions:** The evidence base for population testing lacks high quality studies, however, the literature provides evaluations of the advantages and limitations of different testing modalities. There is a need for robust evidence in this area to ensure that testing is deployed in a safe and effective manner in response to the Covid-19 pandemic.

## Introduction

In response to the SARS-CoV-2 pandemic, many countries implemented population testing programmes as part of countermeasures to contain the spread of infection and mitigate its health and economic impacts. Population testing provides disease surveillance required to inform broader policy decisions, target resource utilisation and, when twinned with timely case isolation and contact tracing, more effective containment of the virus ^1^. Worldwide, population testing programmes are diverse, depending on the population eligible for testing, the technology used to sample and analyse specimens, as well as the timing and frequency of testing.

Population testing seeks to identify people infected with SARS-CoV-2 in a pre-defined group such as health and care workers. By identifying cases of infection through testing, action can be taken to limit infection spread by isolating infected individuals and their contacts during their infectious period. As SARS-CoV-2 may be spread by asymptomatic individuals, including these individuals in testing programmes could help reduce viral transmission. Cumulatively, these actions help control the spread of infection and create conditions that would enable the relatively normal functioning of society.

Modalities used for COVID-19 population testing include drive-in, walk-in, mobile sites, postal testing and home visits. The UK, for example, adopted a testing strategy with 5 pillars, each pillar pertaining to a population subgroup and focusing on either diagnosis, detecting past infection or for surveillance purposes to estimate population prevalence. Testing is co-ordinated centrally and delivered from satellite centres and via postal testing ^2^. South Korea adopted walk-in and drive-through testing ^3, 4^, whilst some areas in Scotland have instituted home-testing to reach more vulnerable groups who cannot access test facilities easily ^5, 6^. There is also increasing political and societal concern of the socioeconomic impact of blunt strategies such as national ‘lockdowns’ to combat COVID-19 ^7^, and their effect on health inequalities. There may therefore be value in studying the different approaches used worldwide, and to learn from successful programmes from other countries.

What is currently unclear is whether any modality of population testing is more robust and efficacious for containing the virus. There is a need to identify population testing programmes that are more accessible and effective at containing the spread of infection, together with the determinants of success. This can help inform national testing policies as part of the pandemic response efforts to minimise the health, social and economic harms. We conducted a scoping review to describe the volume and type of evidence reporting on the experience of implementing population testing for SARS-CoV-2 in high and upper-middle income countries during the pandemic.

## Methods

### Scoping review methodology

Scoping reviews aim to rapidly map the key concepts underpinning a research area, by comprehensively summarising evidence in order to inform practice and policy and provide direction for future research ^8, 9^. Scoping reviews use rigorous and transparent literature searching methods but differ from systematic reviews as the quality of included studies are not routinely assessed, nor do they provide a synthesised answer to a particular research question ^10^.

This scoping review followed the framework proposed by Arksey and O’Malley ^8^ and refined by Levac et al ^11^, briefly comprising: identification of the research question, identification of studies, selection of studies, charting of data and collation of results. This review was commissioned by Public Health England who were consulted on the interim outputs of the study. The trial protocol was published on PROSPERO, number CRD42020186506.

### Identification of the research question

The purpose of this review was to assess the volume of published literature describing the experience of implementing population testing for Covid-19 and to identify the nature and characteristics of the testing programmes. We sought to elucidate what data was available to assess the outcomes of these testing programmes in terms of processes, populations, quality and/or resource-use. We developed these broad aims in order to generate breadth of coverage and map the literature on this topic so that key concepts and gaps could be identified to inform further practice and policy.

Table 1 outlines the inclusion and exclusion criteria for this review. Studies were included if they described the process of providing or accessing a population testing point for symptomatic and/or asymptomatic individuals for Covid-19, using an antigen or antibody test in any setting, using any testing modality. In order to prioritise research relevant to high income countries such as the UK, we included literature relating to comparable health services from high and upper-middle income countries only.

**Table 1.** Inclusion and Exclusion Criteria.

| Inclusion criteria | Exclusion criteria |
| --- | --- |
| • Any study evaluating or describing the process of providing or accessing a population testing point for Covid-19<br>• Symptomatic and/or asymptomatic individuals<br>• Any setting<br>• Antibody or antigen testing<br>• Any testing modality<br>• High or upper-middle income countries according to World Bank criteria | • Commentaries, modelling studies or opinion pieces<br>• Studies describing only laboratory aspects of testing, including studies of diagnostic accuracy.<br>• Studies describing only testing at ports or borders<br>• Studies describing only screening in which samples were not taken<br>• Not written in English |

Studies of laboratory aspects of testing (including diagnostic accuracy), commentaries, opinion pieces and modelling studies were excluded. Studies that described screening where samples were not taken, or that described the testing of passengers at ports or borders, were also considered out of scope and excluded.

### Literature identification and selection

A search strategy (see Appendix 1) was developed to retrieve studies that had evaluated or described the process of providing or accessing a testing point for population testing for Covid-19. An information specialist (AC) searched the electronic databases Medline, Embase and the Cochrane Library (Figure 1). Searches were originally conducted in May 2020 and updated in August 2020. The search was limited to studies in English and published between January and August 2020.

**Figure 1:**
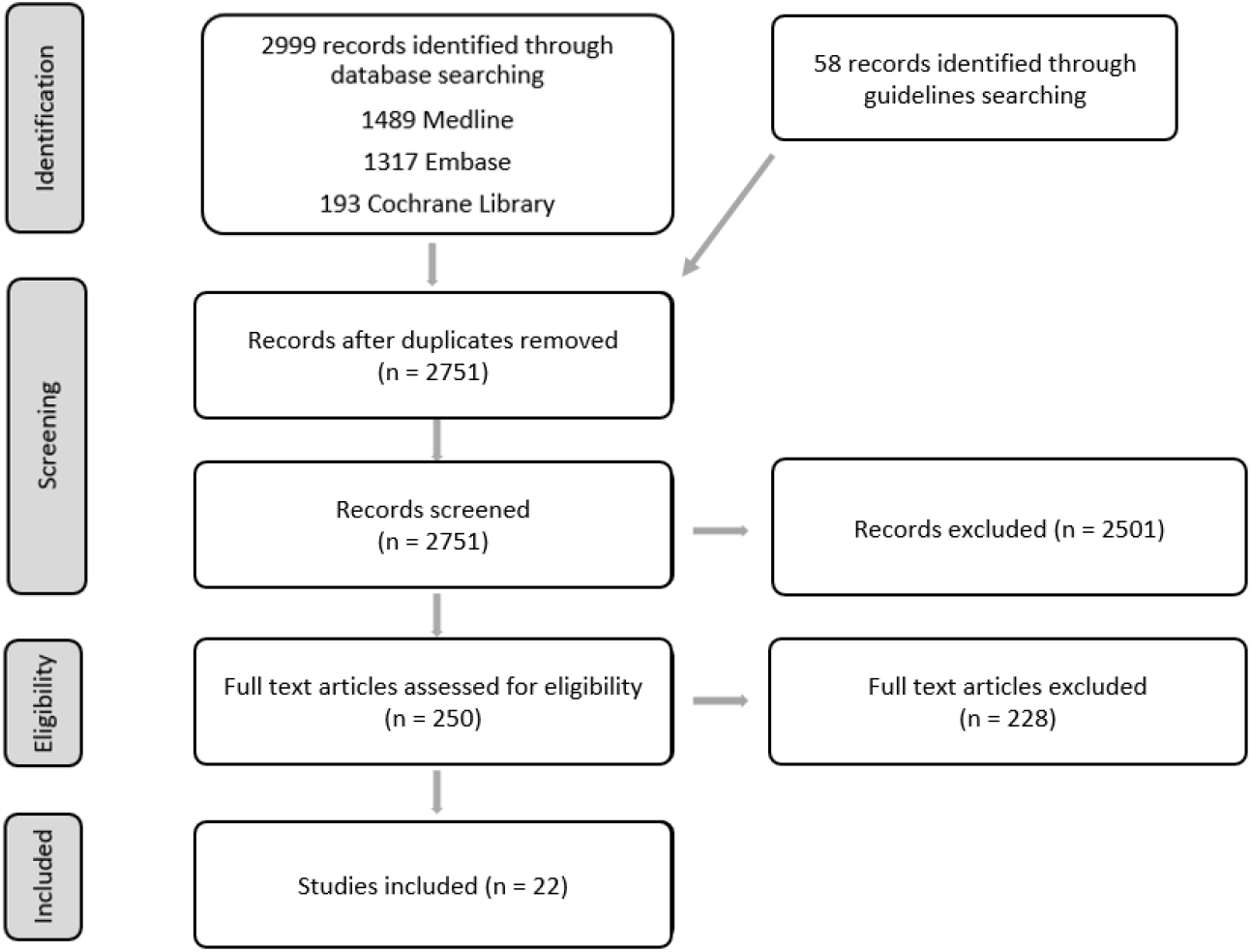
PRISMA flow diagram.

Extracted titles and abstracts were screened by at least two reviewers (CF, FC, LB). 250 full text articles were reviewed to clarify whether the article met the inclusion criteria given in Table 1, either because no abstract was available or because it was unclear from the title and abstract alone whether the study met the inclusion criteria. Abstracts were often unavailable due to the large number of commentaries and opinion pieces found by the search strategy. A formal quality appraisal of the evidence was not conducted, given the remit of the scoping review.

In addition, a grey literature search for national and international clinical guidelines was conducted during May 2020. The World Health Organisation (WHO) and the European Centre for Disease Prevention and Control (ECDC) websites were searched, plus websites in the English language from the UK, USA, Canada, Australia, New Zealand, South Korea, China and Taiwan. This search identified 58 potentially relevant guidelines. On further review, only one guideline was relevant to population testing and detailed an approach to drive-through screening implemented in South Korea ^12^. A further 21 guidelines looked at wider aspects of screening.

### Charting of data and collation of results

After the screening was completed, relevant content in the included studies was extracted into a spreadsheet. Data extraction was verified by a second reviewer who checked data extraction from a random sample of four articles. The mode of testing was categorised into one of five different types: drive-through testing, home visiting testing, indoor walk-though centre, outdoor walk-though centre and mobile testing. For outcome data, a thematic framework was used which categorised any quantitative outcomes indicators into one of four groups: process outcomes, participant outcomes, quality outcomes and resource use outcomes.

## Results

The database search returned 2,999 results (Figure 1). After automated and manual de-duplication, 2,751 unique references were screened for relevance to the question. On first screening of titles, 250 references were identified as potentially relevant, and on further reading 22 were categorised as relevant.

### Study Characteristics

A summary of the studies’ characteristics is presented in Table 2.

**Table 2:**
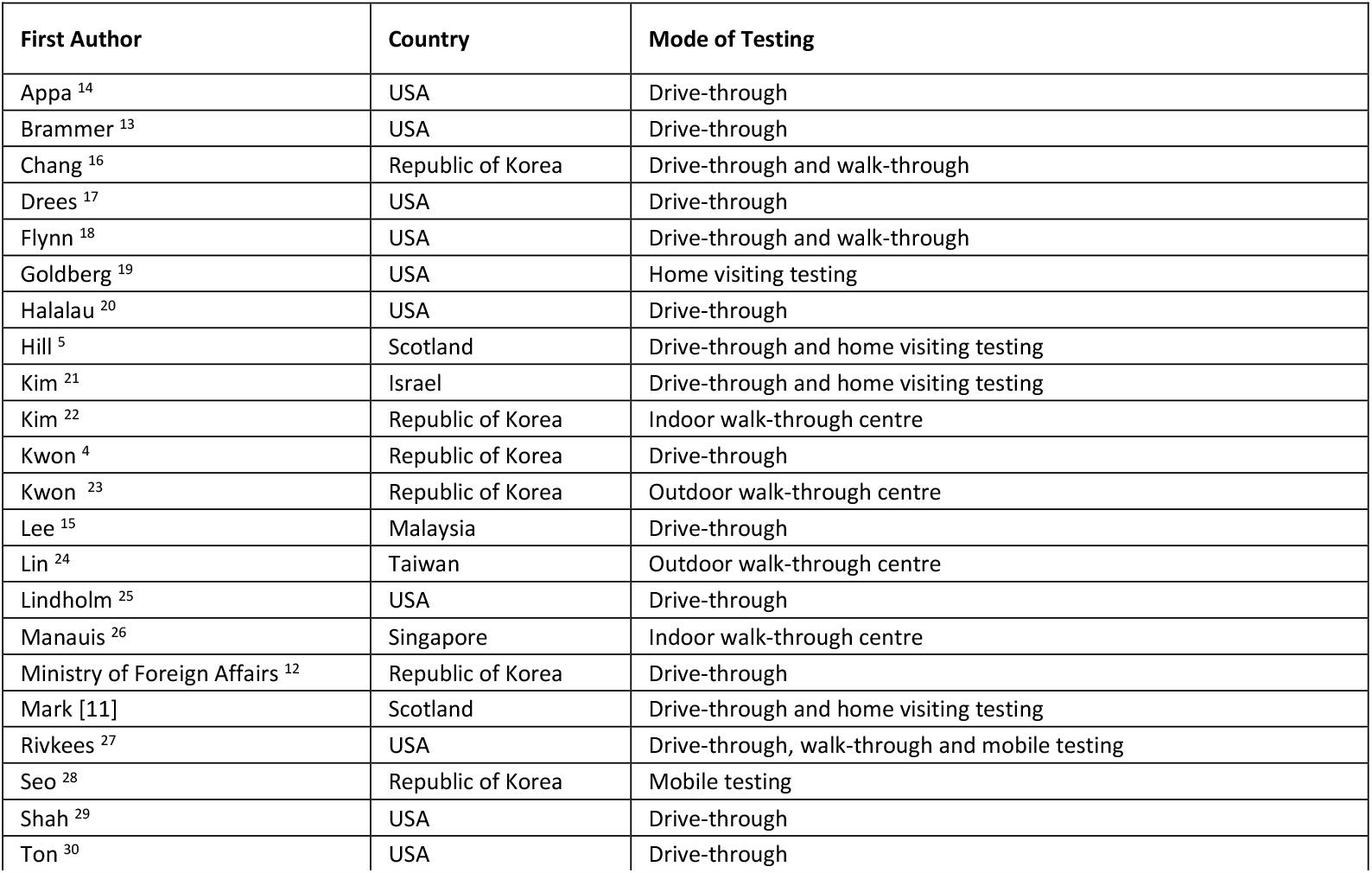
Characteristics of included articles.

| First Author | Country | Mode of Testing |
| --- | --- | --- |
| Appa <sup>14</sup> | USA | Drive-through |
| Brammer <sup>13</sup> | USA | Drive-through |
| Chang <sup>16</sup> | Republic of Korea | Drive-through and walk-through |
| Drees <sup>17</sup> | USA | Drive-through |
| Flynn <sup>18</sup> | USA | Drive-through and walk-through |
| Goldberg <sup>19</sup> | USA | Home visiting testing |
| Halalau <sup>20</sup> | USA | Drive-through |
| Hill <sup>5</sup> | Scotland | Drive-through and home visiting testing |
| Kim <sup>21</sup> | Israel | Drive-through and home visiting testing |
| Kim <sup>22</sup> | Republic of Korea | Indoor walk-through centre |
| Kwon <sup>4</sup> | Republic of Korea | Drive-through |
| Kwon <sup>23</sup> | Republic of Korea | Outdoor walk-through centre |
| Lee <sup>15</sup> | Malaysia | Drive-through |
| Lin <sup>24</sup> | Taiwan | Outdoor walk-through centre |
| Lindholm <sup>25</sup> | USA | Drive-through |
| Manauis <sup>26</sup> | Singapore | Indoor walk-through centre |
| Ministry of Foreign Affairs <sup>12</sup> | Republic of Korea | Drive-through |
| Mark [11] | Scotland | Drive-through and home visiting testing |
| Rivkees <sup>27</sup> | USA | Drive-through, walk-through and mobile testing |
| Seo <sup>28</sup> | Republic of Korea | Mobile testing |
| Shah <sup>29</sup> | USA | Drive-through |
| Ton <sup>30</sup> | USA | Drive-through |

Most of the published literature on this topic is difficult to assign to a study type. Studies were often referred to as brief reports or short communications. The articles typically comprise a description of the testing modality of interest, often with a diagram of the layout of the testing centre and useful operational details followed by an evaluation of its advantages and disadvantages. In some papers, a comparison group was described, however there were no published randomised controlled trials. One article was a qualitative interview study of early experiences of drive-through testing centres ^13^.

All the eligible studies described population testing programmes where samples were taken from symptomatic and occasionally asymptomatic individuals. There were no eligible studies of mass testing (defined as regular and/or large-scale testing of individuals from defined populations regardless of symptom status) that used cheaper, faster tests than Q-PCR. For example, Appa et al. described high throughput, community-wide ascertainment of SARS-CoV-2 prevalence using PCR and antibody testing but not rapid methods such as lateral flow tests ^14^. It was sometimes difficult to categorise articles as ‘mass testing’ because many studies did not describe the eligible population or tests used in detail. It was also often unclear why asymptomatic individuals were attending for testing.

Many articles describing testing programmes were from the USA (43% of studies) and the Republic of Korea (29% of studies). Most articles described testing programmes in high income countries (95% of articles), with only one originating from an upper-middle income country (Malaysia) ^15^.

### Testing Modalities

Several different testing modalities were described, which were categorised into five main categories: drive-through, home visiting, mobile testing, indoor walk-through centres and outdoor walk-though centres.

Sometimes described as off-site COVID-19 testing centres (OSCTCs) ^13^, drive-through testing centres were by far the most common testing modality evaluated (in 72% of articles), in Israel, Malaysia, Korea, Scotland and the USA. This testing modality enabled the use of a vehicle as a self-contained unit, which can reduce the spread of infection. Most were in car parks, stadiums and parks, and one was in an open-air area of a hospital [26]. Some centres enabled individuals without a car to walk in for testing ^13^ in order to increase accessibility. Six articles described drive-through testing in combination with either home visiting testing, mobile testing and/or walk-through testing, enabling a greater proportion of the population to access testing.

Home visiting testing (18% of articles) typically involved a small number of healthcare workers visiting the home of an individual to perform a test. This enabled individuals who are home-bound, frail or have no means of private transportation to access testing without having to use an ambulance, visit a hospital or rely on the assistance of others to access a drive-through site. These schemes were often used as an alternative to local drive-through testing facilities. Home visiting testing took place in Israel, Scotland and the USA.

Two articles described mobile testing. In Korea, testing staff visited workers onsite at their workplaces ^28^ and in Florida, mobile PCR testing laboratories were used to provide point-of-care testing in different cities ^27^.

Indoor walk-through centres based in healthcare facilities were used in Singapore and Korea. There were several different designs for walk-through centres, which were located inside hospitals or other healthcare facilities: screening centres ^22, 26^, negative pressure booths ^22^ and negative pressure tents ^16^. Screening centres permit individuals to access testing inside a building. The Singapore Screening Centre was designed to minimise the movement of patients around the building ^26^. Patients were assigned a seat number and tagged with a tracker to facilitate contact tracing; staff visited patients in their seats to further reduce contact amongst patients.

Negative pressure booths and tents have been designed to minimise the opportunity for viral spread in an indoor setting. Negative pressure booths ^22^ were used for sample collection and medical examination procedures in Korea. The booths were inspired by the design of biosafety cabinets and contain a ‘glove wall’ separating the patient and the medical staff member, who communicate using an interphone. Patients complete registration, questionnaires and payment outside the booths in other sections of the screening centre. Negative pressure booth systems aimed to protect healthcare staff, reduce PPE use and increase throughput compared to other walk-through systems. Negative pressure tents ^16^, also located in Korea, were similar to negative pressure booths, but the whole tent is under negative pressure. Staff working in the tent wore full PPE and most of the tent required sterilisation between people tested that took at least 30 minutes.

The final testing modality described was outdoor walk-through centres. These were located outside hospitals in Korea and Taiwan ^23, 24^. The Korean clinic ^23^ screened all patients and visitors to the hospital with the aim of minimising ward closures due to COVID-19 outbreaks. In Taiwan, a ‘multifunctional sampling station’ was built outside an emergency department, using a 2cm thick clear acrylic board to separate emergency department patients and medical personnel, with inbuilt gloves used to conduct sampling ^24^.

### Populations Tested

Table 3 summarises details of the populations tested and the types of test used. 55% of studies provided information regarding the population eligible to be tested. 41% of articles described accepting both symptomatic and asymptomatic individuals and 14% accepted those with symptoms only. Some testing centres had an algorithm for testing eligibility involving symptoms, epidemiological links, occupational risk factors and/or potential for community exposure. For some, the testing criteria changed over time as the pandemic progressed ^20, 29^.

**Table 3:** Populations eligible to be tested and types of testing used.

| First Author | Population tested | Type of test |
| --- | --- | --- |
| Appa <sup>14</sup> | Symptomatic and asymptomatic | Oro-pharyngeal/mid-turbinate swab for PCR testing, finger prick blood test for antibody testing |
| Brammer <sup>13</sup> | N/A | N/A |
| Chang <sup>16</sup> | Symptomatic and asymptomatic | RT-PCR tests of 'specimens' |
| Drees <sup>17</sup> | Symptomatic only | Not specified |
| Flynn <sup>18</sup> | Unclear/unspecified | 'In-house COVID-19 test' of nasopharyngeal swabs |
| Goldberg <sup>19</sup> | Symptomatic and/or epidemiological link and/or risk factors and/or potential for community exposure | Nasopharyngeal swab collection for RT-PCR analysis |
| Halalau <sup>20</sup> | Unclear/unspecified | Nasopharyngeal swab collection for RT-PCR analysis |
| Hill <sup>5</sup> | Unclear/unspecified | Combined nose and throat swab |
| Kim <sup>21</sup> | Symptomatic only | Swab testing |
| Kim <sup>22</sup> | Symptomatic only | Not specified |
| Kwon <sup>4</sup> | Symptomatic and asymptomatic | Naso- and oro-pharyngeal swabs, sputum specimen |
| Kwon <sup>23</sup> | Symptomatic and/or epidemiological link and/or risk factors and/or potential for community exposure | Pre-testing questionnaire |
| Lee <sup>15</sup> | Symptomatic and asymptomatic | Temperature measurements, nasopharyngeal and oral swabs |
| Lin <sup>24</sup> | Unclear/unspecified | Nasopharyngeal and oral swab, sputum collection, blood testing for antibodies |
| Lindholm <sup>25</sup> | Symptomatic and/or epidemiological link and/or risk factors and/or potential for community exposure | Screening questionnaire followed by nasopharyngeal swab collection for RT-PCR analysis for those with symptoms or epidemiological link |
| Manauis <sup>26</sup> | Symptomatic and/or epidemiological link and/or risk factors and/or potential for community exposure | Swab testing |
| Mark [11] | Unclear/unspecified | Not specified |
| Ministry of Foreign Affairs <sup>12</sup> | Unclear/unspecified | Upper respiratory tract sample (all), lower respiratory tract sample (only if participant can expectorate sputum alone into a container) |
| Rivkees <sup>27</sup> | Symptomatic and asymptomatic | PCR and antibody testing |
| Seo <sup>28</sup> | Unclear/unspecified | Questionnaire followed by sample collection for RT-PCR testing |
| Shah <sup>29</sup> | Symptomatic and/or epidemiological link and/or risk factors and/or potential for community exposure | Originally naso- and oro-pharyngeal swab, later solely nasopharyngeal swab for RT-PCR testing (following CDC advice) |
| Ton <sup>30</sup> | Unclear/unspecified | Nasal swab |

### Types of testing used

The most common method for sampling was through nasal or throat swabs. 45% of articles described the use of nasal swabbing, involving either nasal, nasopharyngeal or mid-turbinate sampling. 27% described the use of throat/oropharyngeal swabbing. 27% stated that swabbing was used but did not specify whether these were nasal and/or throat swabs. Less commonly described testing procedures included sputum sampling (14% of articles) and blood sampling for antibody testing (14% of articles) and one included the use of temperature measurements. 9% of publications described the use of pre-screening questionnaires before sampling took place. 18% of articles did not specify the method of testing and 41% of articles described the use of a combination of the above methods.

### Outcomes of interest

Outcomes of interest were divided into three categories: process outcomes, quality outcomes and resource use outcomes. No participant outcomes were measured quantitatively in any of the included studies, however the discussion section of many papers contained rich qualitative data describing participant outcomes such as staff and participant safety and wellbeing and service equity.

### Process Outcomes

The process outcomes described comprise throughput, duration of test, decontamination time, time to don/doff PPE and waiting time. 77% of articles reported the number of people tested in a specified time period (Table 4). These figures were used to calculate the mean number of people tested each day.^1^ Drive-through testing centres tested between 22-539 individuals per day. Indoor walk-through centres tested 9-500 people per day. One outdoor walk-through centre tested 300 people per day. Home visiting testing teams tested 6-15 people per day.

**Table 4:** Number of people tested per day.

| Number tested per day (calculated) | Pre-screening<br>(Questionnaire/Temperature<br>only) | Testing (Samples taken) | Ref |
| --- | --- | --- | --- |
| Drive-through |  | 22* | 6 |
|  | 107 | 45 | 20 |
|  |  | 60 | 12 |
|  |  | 192 | 30 |
|  |  | >100 | 4 |
|  | 163 | 122 | 25 |
|  |  | 200 (max 400) | 29 |
|  |  | 242 | 16 |
|  |  | 460 | 14 |
|  |  | 539 | 17 |
| Drive-through and home visiting |  | 2,000 country-wide | 21 |
| Drive-through and Walk-through |  | 65 (range 11-127) | 18 |
| Indoor Walk-through centres |  | 9-10 (no negative<br>pressure booths)<br>>70 (with negative<br>pressure booths) | 22 |
|  |  | 41 | 16 |
|  |  | 50-500 | 26 |
| Home visiting testing |  | 6 (max 11)* | 6 |
|  |  | 15 | 5 |
|  |  | 15 | 19 |
| Outdoor Walk-through centre |  | 300 | 23 |

<sup>1</sup> \*2 studies reported the time period per 'week' but did not state which days of the week were available for testing; a 7-day week was assumed for calculations

Some studies compared the throughput of different testing modalities. For similar settings, a higher throughput of individuals could be achieved in a drive-through setting compared to walk-through testing ^16^ or home visiting testing ^5, 6^. For the same time period, one indoor walk-through screening centre using negative pressure booths tested more patients than a walk-through centre with no negative pressure booths (>70 people per day compared to 9-10 people per day) ^22^. Multiple booths could be installed and decontamination time between individuals could be reduced to 3-5 minutes from over 30 minutes. It is difficult to compare different studies, as several variables other than the testing modality can affect the number of people tested per day, such as the number of staff present, the procedures used and the number of individuals who could be tested in parallel.

The mean duration of a drive-through test was between 3-15 minutes. One study reported a median time per test of 28 minutes (IQR 17-44 minutes) ^20^. Some centres allowed multiple people to be tested per vehicle, whereas others allowed only one person per vehicle. The layout of drive-through testing sites can allow several individuals to be tested at one time, for example one drive- and walk-through centre could test two patients every five minutes ^18^.

27% of articles compared testing modalities, calculating a range of indicators for process outcomes (Table 5). Drive-through testing was found to be faster than walk-through testing using a negative pressure tent ^16^ or a screening centre ^4^. Testing using an outdoor walk-through centre (2 minutes per test) was faster than using traditional sample collection in a single negative-pressure isolation room (5 minutes per test) ^24^. Home visiting testing (30 minutes per test) was also quicker than transporting patients to hospital for tests with a specialist ambulance (<1 hour) ^6^. However, waiting times, defined as the time between arrival at the drive-through centre and testing, was reported to be as high as 7 hours at peak volume ^20^.

**Table 5:**
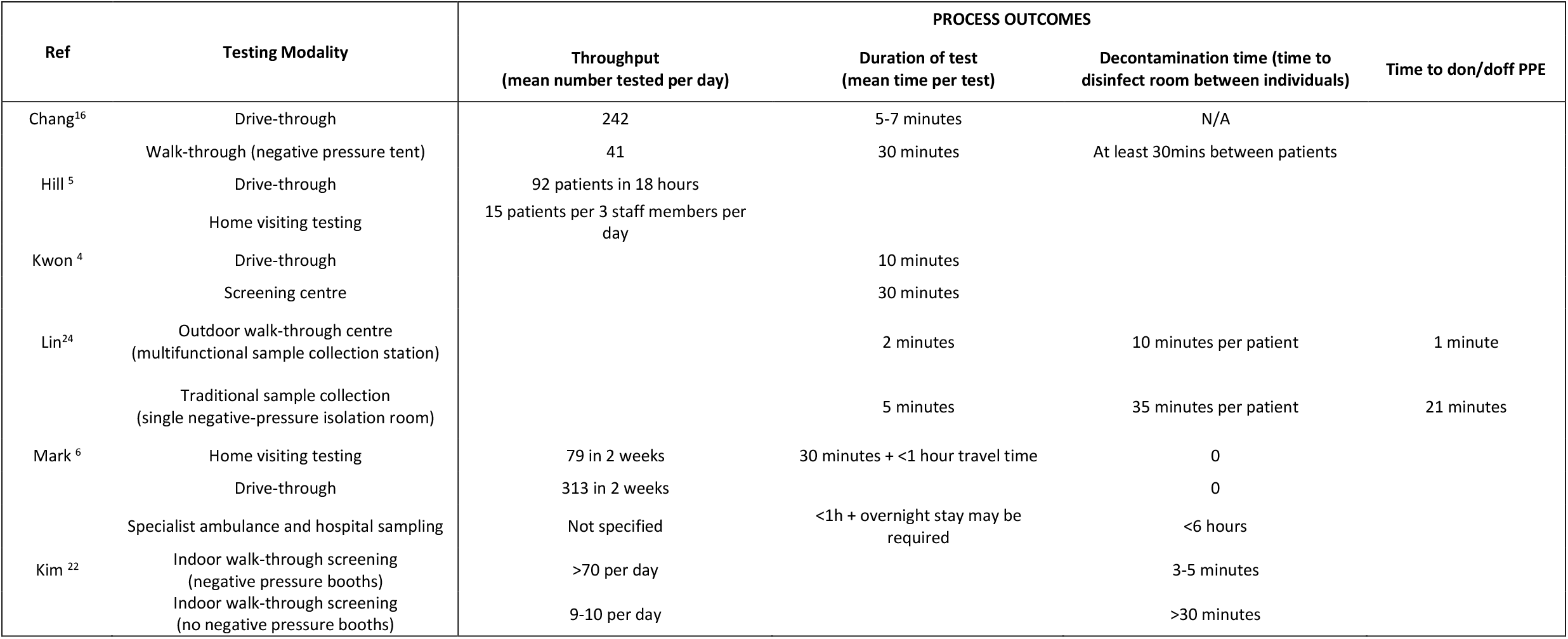
Differences in process outcomes when comparing different testing modalities.

Testing in an outdoor walk-through centre dramatically reduced time to don/doff PPE compared to traditional sample collection in a single negative-pressure isolation room (1 minute per patient compared to 21 minutes per patient) ^24^.

Several studies measured the time to disinfect equipment between individuals (decontamination time). The use of drive-through testing eliminated the need for a 30 minute decontamination time between patients in a walk-through centre using a negative pressure tent ^16^. Drive-through and home visiting testing also required no decontamination time, compared to up to 6 hours of decontamination time needed if a patients was tested in the emergency department ^6^.

Decontamination time was much shorter when using an outdoor walk-through centre (10 minutes per patient) compared to a single negative-pressure isolation room (35 minutes per patient). The use of negative pressure booths reduced decontamination time from >30 minutes with no negative pressure booths to 3-5 minutes between patients ^31^.

### Quality Outcomes

Two different indicators were used to describe quality outcomes: the median time from referral to test and the test turnaround time. Only one study of home visiting testing calculated the median time from referral to test, which was 1 day with a maximum of 3 days ^6^. One drive-through testing study calculated the test turnaround time, defined as the time between testing and communication of results. This was found to be 25 hours (IQR 21-29) in-house and 221 hours (IQR 161-269) if outsourced ^25^.

### Resource Use Outcomes

Resource use outcomes were measured using cost per patient, use of PPE and impact on hospital closure. Home visiting testing reportedly costed much less (£55 per patient) than the use of a specialist ambulance and hospital sampling (£768 per patient) ^6^ (Table 6).

**Table 6:** Resource use outcomes.

| Ref | Testing Modality | RESOURCE USE OUTCOMES |  |
| --- | --- | --- | --- |
|  |  | Cost per patient | PPE use |
| Lin <sup>24</sup> | Outdoor walk-through centre<br>(multifunctional sample collection station) |  | 0 PPE items used* |
|  | Traditional sample collection<br>(single negative-pressure isolation room) |  | 24 PPE items used |
| Mark <sup>6</sup> | Home visiting testing | £55 |  |
|  | Drive-through | Not specified |  |
|  | Specialist ambulance and hospital sampling | £768 |  |
| Ton <sup>30</sup> | Drive-through |  | 1,152 masks, 960 gowns/pairs of gloves for 192 patients |
|  | Emergency department testing |  | 42 masks, 24 gowns, 504 pairs of gloves for 192 patients |

Another study reported that staff in an outdoor walk-through centre used fewer items of PPE than staff working in negative-pressure isolation rooms ^24^. Similarly, drive-through testing can reduce PPE use (96% reduction in mask use, 97% reduction in gown use and 47% reduction in glove use) compared to emergency department based testing ^30^.

One study reported the effect of a screening and testing clinic on maintaining the functioning of a tertiary hospital ^23^. Before the clinic was opened, an average of 36 beds per day were closed due to Covid-19 patients entering the hospital, whereas after the clinic was open and operating well, there was only one closure event (25 beds).

## Discussion

This scoping review provides an overview of the literature describing the experience of implementing population testing for SARS-CoV-2 in high and upper-middle income countries. Whilst a range of modalities were reported, the most commonly evaluated were drive-through services using naso- and/oro-pharyngeal swabbing. Drive-through testing provided a rapid and scalable method of testing for COVID-19, reducing the risk of exposure to staff and patients within healthcare settings and minimising PPE use. However, this approach raises questions regarding equity of access for those who do not have access to a private vehicle or are not well enough to drive. The addition of other testing options such as home visiting, mobile testing or walk-through services may help address this issue.

However, the evidence base for population testing lacks robust studies. Many were simply an evaluation of a testing programme with a discussion of its advantages and limitations rather than robust research studies with control groups. The heterogeneous nature of the testing programmes described in the literature makes it difficult to compare between studies. No studies reported quantitative data on participant outcomes.

There is a paucity of published literature on the implementation of mass testing for SARS-CoV-2. There were no studies on mass testing that fit the inclusion criteria of this review, although a few papers have described mass testing of residents in facilities such as care homes and prisons ^32, 33^. A recent review by the European Centre for Disease Prevention and Control highlights the need for further studies on the assessment and impact of mass testing ^34^. Indeed, some countries have rushed to adopt mass testing before the benefits, risks and costs of this approach is fully understood ^35^. It is therefore pertinent to draw on the international literature on population testing to inform decision making in order to ensure that testing is deployed in a safe and effective way as part of the overall COVID response ^36^.

A distinction needs to be made between population testing and screening for Covid-19. As Covid-19 is a new disease, it is unsurprising that definitions for screening and testing in this context have not yet been standardised and often appear to be arbitrary. We found the term ‘screening’ is loosely used in different ways in the literature, including the testing of symptomatic and/or asymptomatic individuals, assessment of risk factors via a questionnaire, and temperature measurement of individuals travelling past a screening post ^37-39^.

Our literature search aimed to be comprehensive but timely and expedient. Whilst we used rigorous and transparent search methods, we had to limit our search to articles published in English. Therefore it is possible that some relevant studies have not been included. Although we did not formally assess the quality of included papers, much of the literature was not robust as stated earlier. There was also considerable heterogeneity between studies, which limits our ability to compare outcomes from different articles.

Further exploration is needed of population testing using different SARS-CoV-2 tests as the strengths and limitations of the various SARS-CoV-2 tests could influence the yield, cost-effectiveness and viability of the population testing programmes. Additionally, research into the wider consequences of testing programmes is needed especially on population behaviours as a result of testing. Finally, further study of the cost-effectiveness of population testing compared to other pandemic control measures is also required. As with most public health interventions, there is no ‘one-size-fits-all’ approach, and whatever population testing approach is adopted it will need to be tailored to the local context and target population.

## Data Availability

N/A

## Acknowledgements

This work was supported by funding from Public Health England.

## Supplementary Information

**Table 1:** Average time per test.

| Test modality | Mean time per test | Ref |
| --- | --- | --- |
| Drive-through | 10 min per individual (1/3 shorter than conventional testing) | 4 |
|  | 15min appointments per household scheduled (max 5 per car) | 14 |
|  | 5-8 minutes | 15 |
|  | 28 minutes (median) (IQR 17-44). | 20 |
|  | 5-7 minutes | 16 |
|  | 6 lanes of cars in parallel, 3-5min per car | 21 |
| Drive-through and Walk-through | 2 patients swabbed every 5 minutes | 18 |
| Indoor Walk-through centre | 30 minutes | 16 |
|  | 5 minutes | 24 |
| Outdoor Walk-through centre | 2 minutes | 24 |
| Home visiting testing | 30 minutes | 6 |

